# Obesity-Induced Inflammation and the Risk of Alzheimer’s Disease: A Systematic Review

**DOI:** 10.1101/2025.09.18.25336114

**Authors:** Srirachana Reddy Gumireddy, Mariette Anto, Israa Elkashif, Sindhu Vithayathil, Maryam Walizada, Gowtham Siddi, Summayya Anwar

## Abstract

Alzheimer’s Disease(AD) is a neurodegenerative disorder marked by the accumulation of amyloid plaques and tau neurofibrillary tangles in the brain. It is known to be caused by a variety of risk factors, both modifiable and non-modifiable. This systematic review investigates obesity as a modifiable risk factor for AD, focusing on inflammatory pathways linking the two conditions. This review was performed following the Preferred Reporting Items for Systematic Reviews and Meta-Analyses (PRISMA) 2020 guidelines. We searched five databases (PubMed, Cochrane Library, Scopus, ScienceDirect, and Directory of Open Access Journals (DOAJ)) for human studies published in English language in the last 10 years. Following full text analysis and quality assessment, 23 studies were included in the review. We found that obesity induces a state of neuroinflammation through blood-brain barrier and choroid plexus disruptions, adipokine dysregulation, and mitochondrial dysfunction, leading to the development or progression of AD. We also identified nine proteins namely, CHI3L1, PTP1B, GDF15, MMP9, PECAM1, C3AR1, IL1R1, PPARGC1α, and COQ3 as potential biomarkers that may serve as therapeutic targets to delay AD onset or progression. These findings underscore the importance of targeting obesity-related inflammation in AD prevention strategies.

## 1. INTRODUCTION

Dementia is a syndrome that can be caused by a number of diseases which over time destroy nerve cells and damage the brain, typically leading to deterioration in cognitive function beyond what might be expected from the usual consequences of biological ageing. In 2021, 57 million people had dementia worldwide, over 60% of whom live in low-and middle-income countries. Every year, there are nearly 10 million new cases. Alzheimer’s disease is the most common form and may contribute to 60–70% of cases[1].

Alzheimer’s disease is thought to be caused by the abnormal build-up of proteins in and around brain cells. One of the proteins involved is called amyloid, deposits of which form plaques around brain cells. The other protein is called tau, deposits of which form tangles within brain cells[2]. According to data from the Centers for Disease Control and Prevention (CDC), 122,019 people died from Alzheimer’s disease in 2018, the latest year for which data are available[3]. A third of Alzheimer’s disease cases worldwide might be attributable to potentially modifiable risk factors[4]. The modifiable risk factors include low educational attainment, hypertension, hearing impairment, smoking, obesity, depression, physical inactivity, diabetes, low social contact, excessive alcohol consumption, traumatic brain injury (TBI), and air pollution[5].

More recently, growing attention is being given to the impact of obesity on CNS function, as accumulating evidence indicates a higher incidence of neurological disorders in the obese population[6]. Adipocytes release numerous hormones, inflammatory mediators, and immune system effectors into the bloodstream[7]. The expansion and dysregulation of adipose tissue leads to phenotypic shifts in its cellular populations, thereby establishing a chronic low-grade inflammatory environment that is characteristic of obesity [8]. The role of inflammatory pathways associated with obesity in causing neurodegeneration is currently understudied.

This review aims to systematically evaluate the evidence for an association between obesity-induced inflammation and the risk of developing Alzheimer’s disease in adults.

## 2. METHODS

### 2.1 Study Design

A meticulous review was done to assess the risk of development of Alzheimer’s Disease due to obesity-induced inflammation following the Preferred Reporting Items for Systematic Reviews and Meta-Analyses (PRISMA) recommendations[9].

### 2.2 Search Strategy of Databases

A literature search was conducted across five electronic databases: PubMed, Cochrane Library, Scopus, ScienceDirect, and Directory of Open Access Journals (DOAJ). The search strategy employed combined the keywords: “Alzheimer’s disease”, “obesity”, and “inflammation” using Boolean operators and subject-specific terms to identify relevant studies published within the last 10 years. The search methodology was modified in accordance with the specific criteria of each database, including the use of MeSH terms to find relevant results on PubMed. Table 1 details the search strategy used and the results retrieved before and after the application of suitable filters in each database.

**Table 1.** Search Strategy used for each Database and articles identified.

| Database | Keywords | Search Strategy | Search Result before Filter | Filters | Search Result after Filter |
| --- | --- | --- | --- | --- | --- |
| Pubmed | Alzheimer's, obesity, inflammation | (( "Alzheimer Disease/chemically induced"[Mesh] OR "Alzheimer Disease/complications"[Mesh] OR "Alzheimer Disease/diet therapy"[Mesh] OR "Alzheimer Disease/epidemiology"[Mesh] OR "Alzheimer Disease/etiology"[Mesh] OR "Alzheimer Disease/immunology"[Mesh] OR "Alzheimer Disease/metabolism"[Mesh] OR "Alzheimer Disease/physiopathology"[Mesh] )) AND ("Obesity/blood"[Mesh] OR "Obesity/chemically induced"[Mesh] OR "Obesity/complications"[Mesh] OR "Obesity/diet therapy"[Mesh] OR "Obesity/etiology"[Mesh] OR "Obesity/immunology"[Mesh] OR "Obesity/metabolism"[Mesh] OR "Obesity/pathology"[Mesh] OR "Obesity/physiopathology"[Mesh] ) | 288 | Last 10 years, Humans, English, Middle Aged+Age d:45+ years | 35 |
| ScienceDirect | Alzheimer's, obesity | "Alzheimer's Disease" AND "Obesity Induced Inflammation" | 120 | Last 10 years | 98 |
| SCOPUS | Alzheimer's, obesity | "Alzheimer's Disease" AND "Obesity" AND "Inflammation" | 1674 | Last 10 years,<br>Filter by keyword: Alzheimer disease, human, obesity, inflammation on English | 1260 |
| Cochrane | Alzheimer's, obesity | "Alzheimer's disease" AND "Obesity" | 117 | Last 10 years, English | 94 |
| DOAJ | Alzheimer's, obesity | "Alzheimer's Disease" AND "Obesity" AND "Inflammation" | 116 | Last 10 years | 110 |

### 2.3 Inclusion and Exclusion Criteria

Studies were eligible for inclusion if they investigated the potential association between Alzheimer’s disease, obesity, and inflammation. Only studies involving human populations and articles published in English were considered. The patient population under study included adult patients ≥40 years of age with overweight/obesity or obesity as a component of metabolic syndrome. The intervention was inflammation induced by obesity. The primary outcome was the development of Alzheimer’s disease. Exclusion criteria included papers published over 10 years, pediatric and adolescent studies, and animal studies. No restrictions were placed on study design; all types of empirical research studies were considered, including randomized controlled trials (RCTs), cohort studies, case-control studies, cross-sectional studies, and other relevant observational or interventional designs. Non-empirical publications such as editorials, opinion pieces, commentaries, and conference abstracts were excluded.

### 2.4 Data Collection and Quality Appraisal

Two researchers independently extracted data based on the inclusion and exclusion criteria. Full-text articles were thoroughly analyzed, with discrepancies resolved through discussion and consensus.

The quality assessment tools used during the process are: Newcastle-Ottawa Scale(NOS) for case-control and cohort studies, Appraisal Tool for Cross-Sectional Studies(AXIS) for cross-sectional studies, Scale for Assessment of Narrative Reviews Appraisal(SANRA) for narrative reviews, and Strengthening the Reporting of Observational Studies in Epidemiology(STROBE)/ Strengthening the Reporting of Genetic Association Studies(STREGA) for Integrative transcriptome-wide association study. Mixed human-animal studies were evaluated solely on human data. The risk of bias was assessed using the appropriate quality assessment tools for each study design group, and findings are shown in Table 2.

**Table 2.** Quality Appraisal done for different groups of studies.

| Study Design | Total (n) | Quality Assessment Tool Used | Low Risk of Bias (n) | Moderate Risk of Bias(n) | High Risk of Bias (n) |
| --- | --- | --- | --- | --- | --- |
| Cohort | 2 | NOS | 1 | 1 | 0 |
| Case-control | 1 | NOS | 1 | 0 | 0 |
| Cross-sectional | 5 | AXIS | 4 | 1 | 0 |
| Integrative transcriptome-wide association study | 1 | STROBE/ STREGA | 0 | 1 | 0 |
| Narrative reviews | 19 | SANRA | 5 | 9 | 5 |

## 3. RESULTS

### 3.1 Study Selection

In total, 1597 records were retrieved by the preliminary search of the five databases. The citations were exported to EndNote, and 35 duplicate records were removed using EndNote’s automated duplicate removal tool and manually. 1562 studies remained for screening. After meticulous screening of titles and abstracts, 1328 papers were deemed ineligible and thus excluded. 234 studies were retrieved for full-text screening. 17 articles were excluded due to inability to retrieve full text. 217 articles were then assessed for eligibility, of which 194 articles were excluded for a variety of reasons as documented in the PRISMA flow diagram[10] below. Finally, 23 articles were included in this systematic review since they satisfied all inclusion criteria and passed the quality appraisal. A PRISMA flow diagram with the results of the literature review is shown in Figure 1.

**Figure 1.**
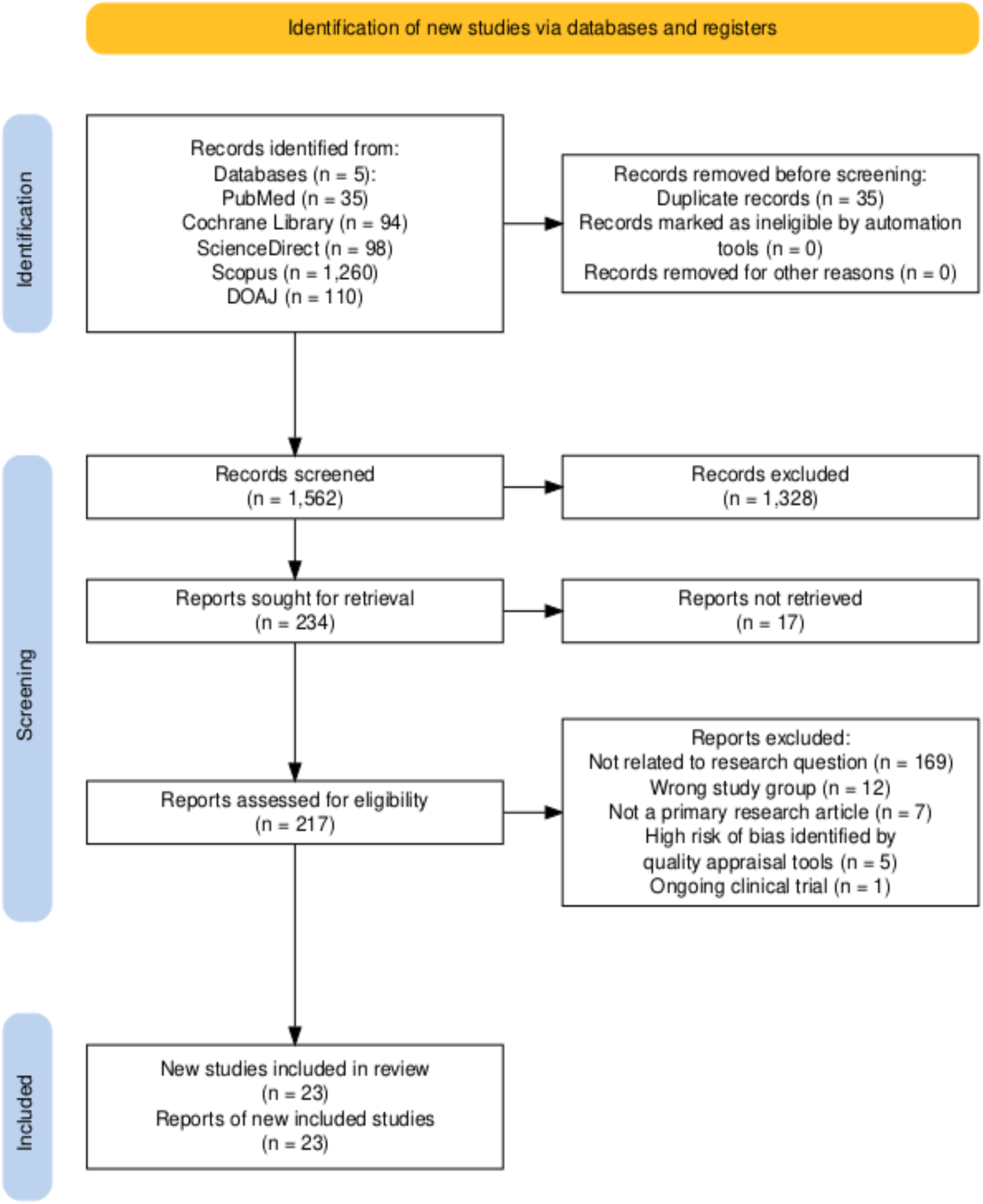
PRISMA Flowchart showing article selection process.

Two cohort studies and a case-control study were appraised using NOS and were included in our study. Similarly, five cross sectional studies were appraised using AXIS and all were included after thorough quality assessment. A combination of STROBE and STREGA was used for a trancriptome wide association study and was scored as having a moderate risk of bias and included in our study. Out of the 19 narrative reviews, 14 were included in our study and 5 were excluded after being scored as having a high risk of bias. This data is depicted in Table 2.

### 3.2 Basal Characteristics of Eligible Studies

A total of 23 studies were included in this review, with the mean age of the study population being 56.4 years. The basal characteristics of the included studies are detailed below in Table 3.

**Table 3.** Basal Characteristics of Eligible Studies.

| Author (Year) | Country | Study Design | Key Conclusion |
| --- | --- | --- | --- |
| Ali et al. (2024)[11] | Germany | Narrative Review | Metabolic syndrome exacerbates Alzheimer's via inflammatory mechanisms. |
| Alisch et al. (2022)[12] | USA | Cross-Sectional | Choroid plexus volume correlates with adiposity in midlife adults. |
| Anagnostakis et al.(2025)[13] | USA | Cross-Sectional | Higher BMI states are associated with faster brain ageing and increased AD-like brain atrophy, particularly in males, while females with normal-weight showed more pronounced brain ageing and AD-like brain atrophy than males with normal-weight males. Second, proteomic analysis revealed 20 proteins postulated to be implicated in the pathophysiological mechanism of brain ageing and AD-like brain atrophy. |
| Bazzari et al.(2023)[14] | Jordan | Narrative Review | Non-genetic risk factors (obesity, hypertension) contribute to dementia. |
| Bednarska-Makaruk et al. (2017)[15] | Poland | Case-Control | Dementia of neurodegenerative origin (Alzheimer's disease and mixed dementia) is characterized by elevated levels of adiponectin |
| Borshchev et al.<br>(2019)[16] | Russia | Narrative Review | Adipokine imbalance is currently viewed as an important mechanism involved in cognitive dysfunction in obesity, while normalization of altered leptin and adiponectin signaling in the brain is emerging as one of the valid targets for prevention and treatment of dementia. |
| Corlier et al.(2018)[17] | USA | Cohort | Midlife metabolic syndrome predicts Alzheimer's pathology progression. |
| Costache et al.(2023)[18] | Romania | Narrative Review | Obesity-related inflammation may be associated with earlier Alzheimer's onset. |
| De A. Boleti et al.(2023)[19] | Brazil | Narrative Review | Obesity, insulin resistance, and dyslipidemia converge in an inflammatory process, neurological dysfunction, and neurodegeneration. |
| Dyken et al. (2018)[20] | Canada | Narrative Review | Metabolic syndrome disrupts blood-brain barrier integrity. |
| Emmerzaal et al.<br>(2015)[21] | USA | Cohort | In midlife, increased BMI has been shown to be a risk factor for dementia and AD, whereas this is not, or to a lesser extent, observed in later life. |
| Fontana et al. (2021)[22] | USA | Narrative Review | Caloric restriction reduces neuroinflammation in adults $\geq 40y$. |
| Forny-Germano et al.<br>(2019)[23] | Brazil | Narrative Review | Dysregulated adiponectin and leptin signaling may mediate the detrimental impact of obesity on CNS and raise the risk for cognitive decline and AD. |
| Ge et al. (2024)[24] | China | Cross-Sectional | PTP1B levels are increased in the forebrain of patients with neurodegenerative disease, and also increased in the serum of obese individuals who have increased white matter and cerebrospinal fluid volumes. White matter abnormalities are considered a core feature of neurodegeneration. |
| Ha et al. (2023)[25] | South Korea | Cross-Sectional | Higher leptin-to-adiponectin ratio(LAR) was significantly associated with CRP and HOMA-IR. In addition, LAR was associated with increased odds of AD, and its estimated risk was higher than that associated with either leptin or adiponectin levels alone. |
| Khan et al. (2020)[26] | USA | Narrative Review | Chronic inflammation bridges obesity-diabetes-Alzheimer's triad. |
| Li et al. (2022)[27] | China | Integrative transcriptome-wide association study | Increased inflammation/immune response and mitochondrial dysfunction in obesity might be an essential susceptible factor for AD. |
| Ly et al. (2021)[28] | USA | Cross-Sectional | This study builds upon a growing understanding of obesity brain dysfunction by establishing a relationship between white matter extracellular water as a proxy of neuroinflammation and obesity as measured by BMI. Additionally, this work also links obesity related white matter edema with volume loss in the hippocampus. |
| Marino et al. (2025)[29] | Italy | Narrative Review | Mitochondrial dysfunction in obesity extends to CNS. |
| Marrano et al. (2023)[30] | Italy | Narrative Review | Cellular lipotoxicity mediates diabetes-Alzheimer's crosstalk. |
| Mekhora et al. (2024)[31] | Thailand | Narrative Review | In the context of humans, either acute or chronic inflammatory status showed an association with cognitive decline. |
| Neto et al. (2023)[32] | Portugal | Narrative Review | Obesity impacts neurodegenerative disorders through shared underlying mechanisms, underscoring its potential as a modifiable risk factor for these diseases. |
| Vinuesa et al. (2021)[33] | Argentina | Narrative Review | Insulin resistance and chronic inflammation are two synergic phenomena underlying the pathogenesis of metabolic diseases as well as the development of the wide variety of associated disorders, including AD-like neurodegeneration. |

## 4. DISCUSSION

### 4.1 Pathophysiology of Alzheimer’s Disease

Alzheimer disease is characterized by gradual and progressive neurodegeneration caused by neuronal cell death. The neurodegenerative process typically begins in the entorhinal cortex within the hippocampus[34].

The 2 pathologic hallmarks of Alzheimer disease are

● Extracellular beta-amyloid deposits (in neuritic plaques)
● Intracellular neurofibrillary tangles (paired helical filaments)

The beta-amyloid deposition and neurofibrillary tangles lead to loss of synapses and neurons, which results in gross atrophy of the affected areas of the brain, typically starting at the mesial temporal lobe. The amyloid hypothesis posits that progressive accumulation of beta-amyloid in the brain triggers a complex cascade of events ending in neuronal cell death, loss of neuronal synapses, and progressive neurotransmitter deficits; all of these effects contribute to the clinical symptoms of dementia[35]. A sustained immune response and inflammation have been observed in the brain of patients with Alzheimer disease. Some experts have proposed that inflammation is the third core pathologic feature of Alzheimer disease[36]. Unlike other risk factors and genetic causes of AD, neuroinflammation is not typically thought to be causal on its own but rather a result of one or more of the other AD pathologies or risk factors associated with AD and serves to increase the severity of the disease by exacerbating β-amyloid and tau pathologies[37],[38].

Prior literature has suggested multiple risk factors that contribute to at least 50% of AD risk[39]. Chief among modifiable risk factors is having excess body tissue adiposity, characterized as being overweight and obese, an increasingly pervasive public health problem projected to affect 85% of the U.S. and 58% of the global population by 2030[40]. The increase in adipocytes in obesity triggers adipocyte differentiation in a process called adipogenesis, in which pro-inflammatory mediators or adipokines are generated (leptin, adiponectin, resistin, TNF-α, IL-1β, –6 and –8, insulin-like growth factor 1, monocyte chemoattractant protein-1, and visfatin)[41]. This systematic review aims to identify evidence of an association between obesity and risk of development or progression of Alzheimer’s disease through inflammatory mechanisms.

### 4.2 Obesity and Alzheimer’s Disease: Changes in Choroid Plexus and Cerebrospinal Fluid

White matter vasogenic edema is a promising marker of neuroinflammation that may offer a mechanism for the effects of obesity on brain structure[42],[43]. A study conducted by Ly et al[28] in 104 participants(42 normal weight controls with mean BMI=22.7±1.9, mean age= 63.0±7.7 years, and 62 overweight or obese individuals with mean BMI=29.9±3.5, mean age= 61.3±7.9 years) aimed to quantify patterns of neuroinflammation within cognitively normal, middle-to older-aged individuals as a function of elevated body mass index (BMI). The study evaluated whether neuroinflammation imaging(NII) derived edema fraction in the white matter tracts varied in individuals as a function of normal or increased BMI. They then related white matter tract neuroinflammatory edema to hippocampal volumes, as well as CSF biomarkers of neurodegeneration and neuroinflammation. The study demonstrated statistically significant relationships between neuroinflammation, elevated BMI, and hippocampal volume, raising implications for neuroinflammation mechanisms of obesity-related brain dysfunction in cognitively normal elderly[28].

Implicated in mediating the neuroinflammatory effects of obesity is the choroid plexus (CP) [44–46], a critical cerebral structure necessary for cerebrospinal fluid (CSF) production. A study conducted by Alisch et al[12] aimed to elucidate the relationship between lateral ventricle(LV) volume, CP volume, and CP microstructure, which was assessed using *T*1, *T*2(longitudinal and transverse relaxation times) or mean diffusivity(MD), and BMI or WC, as measures of obesity. The final cohort consisted of 123 cognitively unimpaired volunteers ranging in age from 21 to 94 years (55.0 ± 20.5 years), of which 65 were men (56.0 ± 21.5 years) and 58 were women (54.0 ± 19.6 years). The cohort consisted of 56 lean participants (BMI < 25), 50 overweight participants (25 ≤ BMI < 30), and 17 participants with obesity. The study found a modest difference in MD values and a significant difference in *T*1 values between obese and lean participants after correcting for age, sex, and ethnicity, with MD and *T*1 exhibiting higher values with WC. Stratification by the BMI cutoff revealed significant differences in T1 and MD between obese and overweight individuals. The study concluded that obesity may represent a modifiable risk factor for disruption of CP structure[12], resulting in neuroinflammation.

### 4.3 Obesity and Alzheimer’s Disease: Changes in Cortical Thickness

Another study by Corlier et al[17] used brain magnetic resonance imaging (MRI) to scan 335 older adult humans (mean age 77.3 ± 3.4 years) who remained non-demented for the duration of the 9-year longitudinal study and measured baseline CRP levels. The study found that higher metabolic risk(derived from body mass index, serum insulin, and plasma triglyceride levels) was associated with higher peripheral CRP, which was in turn associated with thinner cortex[17], a finding suggestive of neurodegenerative diseases like Alzheimer disease.

### 4.4 Biomarkers in Alzheimer’s Disease

A study by Anagnostakis et al[13] sought to evaluate machine learning (ML)-based neuroimaging markers of brain age and AD-like brain atrophy in participants with obesity or overweight without diagnosed cognitive impairment (WODCI), in a study of 46,288 participants in 15 studies (the Imaging-Based Coordinate System for Aging and Neurodegenerative Diseases (iSTAGING) consortium). They also assessed the association between cognition, serum proteins, and brain ageing indices. Of the 46,288 participants, 24,897 were females and 21,391 were males, with a mean age of 64.33 years (SD = 8.13) and a mean BMI of 26.81 kg/m2 (SD = 4.49). The study found that the impact of obesity on brain ageing, and AD-like brain atrophy is more pronounced in males than in females, and its effects are weaker with increasing age. Moreover, higher BMI states are linked to accelerated brain ageing and AD-like brain atrophy, especially in males, whereas females with normal-weight exhibited greater brain ageing and AD-like brain atrophy compared to their male counterparts with normal-weight. Proteomic investigations of two time-points revealed five proteins that are associated positively with both brain ageing and BMI i.e., LY96, GDF15, PIGR, CHI3L1, and OXT[13]. GDF15, a cytokine in the transforming growth factor β superfamily, is linked to decreased brain volume in Alzheimer’s-affected regions[47]. Elevated in inflammatory states, GDF15 poses a high risk for AD-like brain atrophy patterns[48],[49]. Similarly, CHI3L1 is an inflammatory marker increased in obesity and diabetes and related to insulin resistance; it has been frequently investigated in body fluids as a surrogate marker of neuroinflammation in AD[50].

Adipokines are secreted factors which carry regulatory signals from adipose tissue through systemic circulation to control a wide range of physiological functions throughout the human body. Dysregulated adiponectin and leptin signaling may mediate the detrimental impact of obesity on CNS and raise the risk for cognitive decline and AD[23]. A study was conducted by Bednarska-Makaruk et al[15] in 425 subjects. 89 patients with mean age 72.8 ± 8.13 years were diagnosed with probable Alzheimer’s disease and the mean age for 107 age-matched controls was 71.3 ± 7.95 years. Serum adiponectin, leptin and resistin levels were measured. The study found that significantly higher levels of leptin were associated with the presence of abdominal obesity in all investigated groups. The study also demonstrated a positive correlation of leptin with some pro-inflammatory indices IL-6 and hsCRP and negative correlation with HDL cholesterol considered as a negative indicator of inflammation. The study also showed positive correlation of resistin with inflammatory indicators (interleukin-6 and CRP) and a negative correlation with anti-inflammatory markers (HDL-C and PON1 activity) in all dementia patients. The study also found a significantly lower level of adiponectin was associated with the presence of abdominal obesity[15]. This study correlates with a study conducted by Ha et al[25] in which 171 participants were enrolled and divided into participants with obesity and without obesity to explore the effect of obesity on the relationship between adipokines and cognition. All the participants were aged 52–95 and the mean age of participants was 74.30 ± 6.63 years and divided into two groups as without obesity (BMI < 25 kg/m2) and with obesity (BMI ≥ 25 kg/m2). The study found that cognitive function was negatively associated with leptin levels and leptin-to-adiponectin ratio (LAR). Such correlations between leptin and cognitive domains were prominent in participants with obesity but were not observed in those without obesity. Leptin levels were associated with lower hippocampal volumes in participants with obesity. A significant interaction of leptin and obesity was found mostly in the medial temporal lobe. Both leptin and LAR were positively associated with insulin resistance and inflammation markers in all participants. Of note, LAR was associated with a higher risk of AD after adjusting for demographic variables, Apolipoprotein E genotype, and body mass index[25]. Restoring proper leptin and adiponectin signaling in the brain may constitute beneficial, disease-modifying therapeutic interventions[23] in AD.

Protein-tyrosine phosphatase 1B (PTP1B), a key inhibitor of signaling pathways for synaptogenesis, is overexpressed by proinflammatory cytokines, such as TNF-α[51]. PTP1B gene and protein expression are remarkably higher in peripheral metabolic tissues, including liver and adipose tissues of obese, insulin-resistant, or diabetic humans [52], [53], [54], [51]. A study conducted by Ge et al[24] investigated PTP1B levels in the serum and forebrain of obese individuals and neurodegenerative diseases patients, with further analysis of the correlations of PTP1B with cognition. Seventy-one patients with type 2 diabetes, thirty-six obese individuals (BMI = 31.01 ± 2.85 kg/m2) (19 male and 17 female) and thirty-five age– and sex-matched lean controls (BMI = 22.19 ± 1.41 kg/m2) (21 male and 14 female), were recruited. The study reported that PTP1B levels were significantly increased in the serum of obese subjects and PTP1B levels were correlated with cognitive decline accompanied by enhanced white matter and cerebrospinal fluid volumes in obese humans[24]. These changes in the cerebrospinal fluid volumes correlates with the CP volumes in obese individuals in the study conducted by Alisch et al[12].

### 4.5 Obesity and Alzheimer’s Disease: Mitochondrial Dysfunction

Marino et al[29] concluded in their study that although the exact underlying mechanisms remain unclear, it has been hypothesized that the adipose tissue dysfunction that occurs in obesity leads to systemic inflammation and to the alteration of the BBB resulting in neuroinflammation and cognitive decline. In this scenario, mitochondrial dysfunction plays a crucial role[29]. Obesity consistently results in mitochondrial dysfunction[55]. Meanwhile, the mitochondrial impairment of obesity is able to stimulate the production of reactive ROS further to promote inflammation, thereby accelerating Alzheimer’s disease progression[56]. Therefore, bioactive compounds that enhance mitochondrial function and eliminate excess ROS can be used to treat NDDs[29]. Li et al[27] in their study illustrated the possible mechanism of AD secondary to obesity(OB) via novel bioinformatic tools and approaches. It was revealed that the increased inflammation/immune response and mitochondrial dysfunction in OB might be an essential susceptible factor for AD and identified novel gene candidates (MMP9, PECAM1, C3AR1, IL1R1, PPARGC1α, and COQ3) that could be used as biomarkers or as potential therapeutic targets[27].

### 4.6 Obesity and Alzheimer’s Disease: Age Discrepancy

Although many studies show the association between late-life obesity and Alzheimer’s disease, a decade of research (2003–2013) by Emmerzaal et al[21] investigating the association between AD, dementia, and BMI has yielded a general picture of how this risk relationship may change over the life course from mid-to late-life. They concluded that in midlife, increased BMI has been shown to be a risk factor for dementia and AD, whereas this is not, or to a lesser extent, observed in later life[21].

## 5. LIMITATIONS

To maximize inclusivity of relevant evidence, narrative reviews were appraised using a lenient application of the modified SANRA criteria, while primary analysis focused on the 9 included empirical studies. This approach has several limitations which are: The observational nature of most included studies precludes causal conclusions; Heterogeneous definitions of both obesity (varying BMI/WC thresholds) and inflammation markers across studies may affect comparability; and while lenient SANRA evaluation allowed broader consideration of theoretical frameworks from narrative reviews, these were only used to bridge literature gaps and were not included in quantitative synthesis to maintain methodological rigor.

## 6. CONCLUSION

This systematic review elucidates obesity-induced neuroinflammation as a critical pathway in Alzheimer’s disease (AD) pathogenesis. Analysis of 9 empirical studies revealed: Inflammatory markers like CRP, and IL-6 in obesity show stronger associations with AD neuroimaging, suggesting blood-brain barrier disruption as a key mechanism; Adipokine dysregulation (leptin resistance, reduced adiponectin) mediates cognitive decline; Choroid plexus structural changes and CSF proteomic signatures like GDF15, CHI3L1, and PTP1B emerge as promising biomarkers for early AD detection in obese populations; and Mitochondrial dysfunction occurs due to elevated ROS in obesity and novel gene candidates like MMP9, PECAM1, C3AR1, IL1R1, PPARGC1α, and COQ3 could be used as biomarkers in AD and for potential therapeutic targeting. These findings position obesity-related inflammation as a modifiable risk factor and identify targeted anti-inflammatory strategies as potential interventions to delay AD progression. The urgent need for clinical translation is underscored by the growing global burden of both obesity and AD, coupled with the absence of current therapies targeting this mechanistic intersection. Future research should prioritize randomized controlled trials that evaluate whether metabolic interventions can delay AD onset or progression in high-risk obese populations.

## DATA AVAILABILITY STATEMENT

No new data was generated in this study. This is a systematic review of existing published literature.

